# Impact of non-pharmacological interventions on COVID-19 boosting vaccine prioritization and vaccine-induced herd immunity: a population-stratified modelling study

**DOI:** 10.1101/2021.10.27.21265522

**Authors:** Zhiyao Li, JiaLe Wang, Boran Yang, Wenjing Li, Jian-Guo Xu, Tong Wang

## Abstract

**Background:** While the COVID-19 pandemic seemed far from the end, the booster vaccine project was proposed to further reduce the transmission risk and infections. However, handful studies have focused on questions that with limited vaccine capacity ether boosting high-risk workers first or prioritizing susceptible normal individuals is optimal, and vaccinating how many people can lead us to the goal of herd immunity. In this study we aimed to explore the conclusions of such two problems with consideration of non-pharmacological interventions including mandatory quarantine for international entrants, keeping social distance and wearing masks.

**Methods:** By implementing the corresponding proportion of individuals who remain infectious after four lengths of quarantine strategies to the novel population-stratified model, we quantified the impact of such measures on optimizing vaccine prioritization between high-risk workers and normal populations. Furthermore, by setting the hypothetical COVID-19 transmission severity (reproduction number, *R*_0_) to the level of the most contagious COVID-19 variant (B.1.617.2, delta variant, *R*_0_ = 5.0), we separately estimated the threshold vaccine coverage of five countries (China, United States, India, South Africa and Brazil) to reach herd immunity, with and without the consideration of interventions including wearings masks and keeping social distance. At last, the sensitive analysis of essential parameter settings was performed to examine the robustness of conclusions.

**Results:** For Chinese scenarios considered with moderate hypothetical transmission rate (*R*_0_ = 1.15–1.8), prioritizing high-risk workers the booster dose reached lower cumulative infections and deaths if at least 7-days of quarantine for international travelers is maintained, and the required screening time to remain such vaccinating strategy as optimal increased from 7-days to 21-days with the transmission severity. Although simply maintaining at least 7-days quarantine can lead to over 69.12% reduction in total infections, the improvement of longer quarantine strategies was becoming minimum and the least one was 2.28% between the 21 and the 28-days of quarantine. Besides, without the vaccination program, the impact of such measures on transmission control dropped significantly when *R*_0_ exceeded 1.5 and reached its minimal level when *R*_0_ equal to 2.5. On the other hand, when we combat the delta variant, the threshold vaccine coverage of total population to reach herd immunity lay within 74%–89% (corresponding to the vaccine efficiency from 70% to 50%), and such range decreased to 71%–84% if interventions including wearing mask and keeping social distance were implemented. Furthermore, Results of other countries with 85% vaccine efficiency were estimated at 79%, 91%, 94% and 96% for South Africa, Brazil, India and United States respectively.

**Conclusions:** Non-pharmacological interventions can substantially affect booster vaccination prioritization and the threshold condition to reach herd immunity. To combat the delta variant, restrictions need to be integrated with mass vaccination so that can reduce the transmission to the minimum level, and the 21-days might be the suggested maximum quarantine duration according to the cost-effectiveness. Besides, by implementing interventions, the requirement to reach herd immunity can be lower in all countries. Lastly, the following surveillance after vaccination can help ensure the real-time proportion of vaccinated individuals with sufficient protection.

## Background

As the COVID-19 remains the state of spreading globally, emerging variants with higher transmissibility posed serious threat to the human health. Imperfect vaccine efficiency against infections, along with the vaccination hesitation, led the pandemic far from the end[1]. To restore the public order with existing flawed vaccine, administrators consecutively announced the booster dose vaccination project for targeting populations[2,3]. Although it was an appealing idea that further reduce the risk or the number of COVID-19 infections by enhancing the immunity of people with the additional shots[4,5,6], limited evidences discussed the problem that vaccines should be used in boosting individuals who work or live in high-risk settings first or prioritizing normal susceptible ones, and whether non-pharmacological interventions can influence such prioritization strategies.

On the other hand, countries with the goal to eliminate COVID-19 transmission were exhausted to encounter recurrent domestic outbreaks[7,8,9]. While they tried their best to promote people getting vaccinated, non-pharmacological interventions including the quarantine policy for oversea travelers were also being implemented in order to reach the temporarily disease-free state. However, several outbreaks caused by asymptomatic outliers in China which hold the most restrictive quarantine policy of the world indicated that such measures were effective but not enough to end the invasion of virus[10,11], while maintaining such restriction was also a pain in the neck to the economics of the developing country[12]. Thus the question that to what extent do people getting vaccinated combined with restrictions could provide stable protection is yet to be answered.

Age-structured mathematical models proposed in previously published studies successfully simulated the COVID-19 transmission[13,14,15]. Due to a number of polices were made for specific populations or conducted with prioritization, as an extended version, we developed a population-stratified model by further dividing the total populations to two sections in order to better simulate the interactions between them. By using this model, we are able to quantify the impact of non-pharmacological interventions on optimizing vaccine prioritization between boosting high-risk population first or allocating to susceptible individuals of normal population. In this study, we also explored the threshold condition of reaching herd immunity by vaccine- and interventions-induced protection.

## Methods

### Infective outliers distribution

In order to establish reliable quarantine time suited for all infection types, we previously estimated the distribution of COVID-19 incubation period with exceeding 1900 cases collected from public available datasets, including not only common symptomatic infections but also the asymptomatic ones (we used the date of testing positive as that of symptom onset for the latter type)[16]. Thus the infective outliers distribution can be statistically interpreted as the survival function of such incubation period distribution:

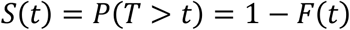

Where the infectious outliers distribution *S*(*t*) is a monotonically decreasing function of incubation(or quarantine) time *t*.

By using bootstrap method, we obtained the mean and essential percentiles of the distribution (In this study we fitted the incubation time into log-normal distribution). The mean value of outliers rates under four different quarantine duration were applied in the proposed mathematical model so that the impact of quarantine measures on COVID-19 transmissions can be accurately quantified.

### Population-stratified model

To characterize the epidemic induced by oversea imported infections under different settings of quarantine time, we applied the population-stratified SEIR (susceptible, exposed, infectious, recovered) model and the total population were divided to two sections: (1) The normal population which was categorized to nine age groups (eight groups for 0-80 years old people per 10 years and one for 80 years old above) due to the varying age demographics in countries and age-specific susceptibility and mortality[17,18]; (2) The high-risk population who contacted infections more frequently due to their occupations (Specific related occupations were listed in Additional file 1). According to the law of closed-loop management for travelers arriving in China, infections identified during the screening period will be transferred from quarantine facilities to hospitals by professionals, thus merely asymptomatic carriers with negative nucleic acid test results throughout the quarantine but remain infectious afterwards had the chance to infected normal individuals (See infections definition in Additional file 1). The sources of infections for two categorized population were illustrated in Fig. 1.

**Fig. 1:**
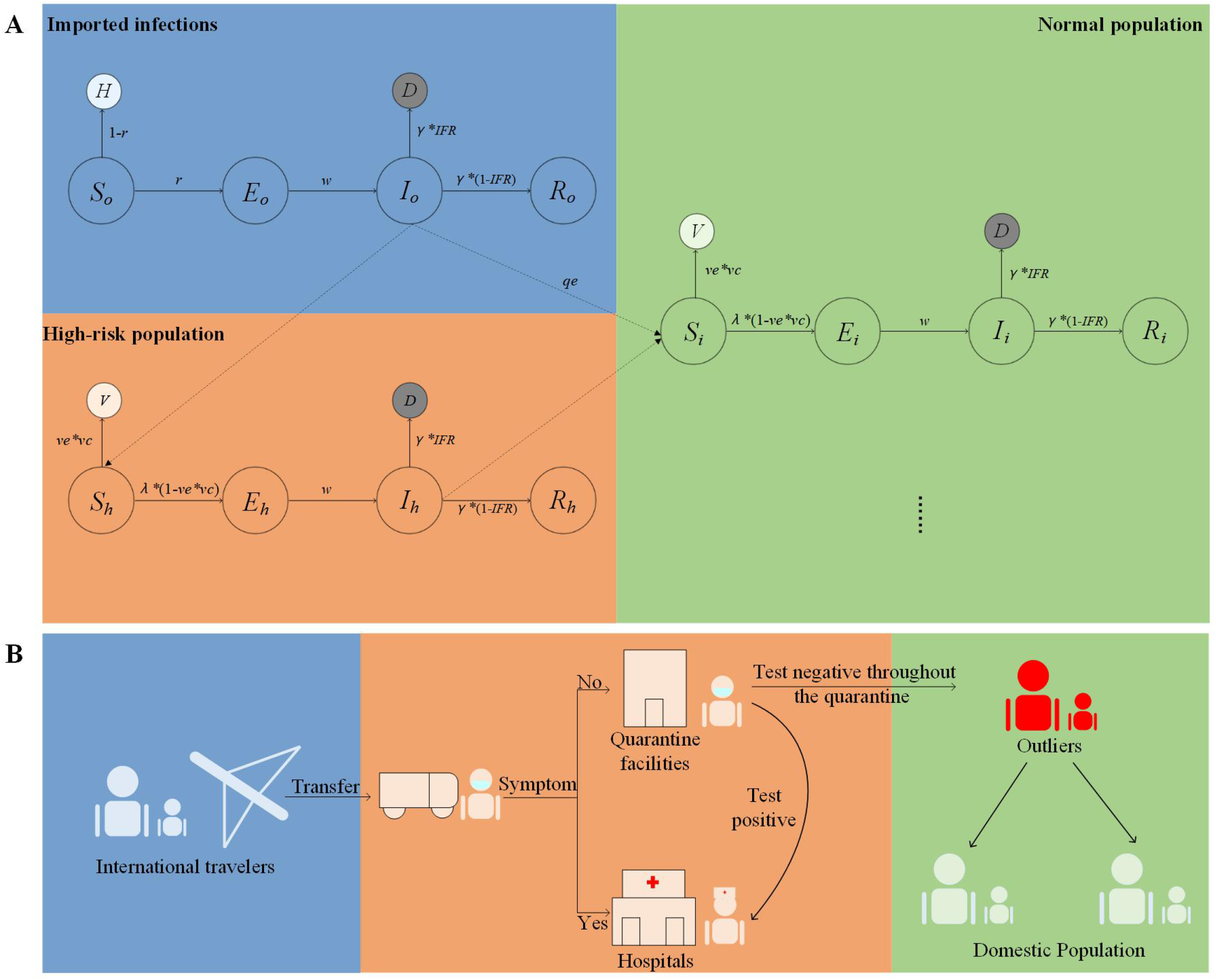
Schematics for the model framework and transmission route. **A** Diagram of the population-stratified model. We divided the total population into two sections: the high-risk workers and the normal population which were further categorized to nine age groups at pace of 10 years. Compartments are susceptible (*S*), exposed (*E*),infected (*I*),recovered (R),vaccinated(*V*) and healthy(*H*). Dots represent the rest eight age groups. **B** The closed-loop management for travelers arriving in mainland China diagram. Merely asymptomatic carriers with negative nucleic acid testing results throughout the quarantine but remain infectious afterwards have the chance to infected normal people.

In this study we assumed that the high-risk population always held higher levels of contacts with infections than normal ones, including the scenario without quarantine settings. We also made the assumption that the type of vaccine efficiency (*ve*) considered in this model was all-or-nothing, which meant from the probabilistic perspective, the vaccine provide perfect protection for a fraction of people (*ve*) once they receive the shot[19]. Lastly, for countries with existing vaccine coverage, as the vaccination-induced neutralizing antibody (NAb) levels decreased with time, we assumed that 50% of already vaccinated individuals were immune to infections and thus the rest remained susceptible, which were considered to be qualified for vaccination in the simulation[20,21,22]. Other related assumptions that might affect the result were described in Additional file 1.

We used the next generation method to obtain the basic reproduction number *R*_0_, which was the absolute value of the dominant eigenvalue of matrix *M*:

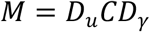

The *D*_*u*_ and *D*_*y*_ represented the diagonal matrix of susceptibility and infectious period respectively, and *C* was the country-specific contact matrix. By calculating the scaling constant of the targeting *R*_0_, we were able to simulate different transmission severity or variants, which can be mathematically interpreted as the change of original contact rates. Such settings were implemented prior to the simulation.

### Boosting vaccine prioritization

We first used the proposed model with one-year simulation to ascertain the optimal vaccine prioritization between boosting high-risk workers first or prioritizing susceptible normal individuals under different quarantine duration and transmission severity (*R*_0_) settings. Firstly, the best vaccinating strategies that reached lower infections and deaths under three lengths of surveillance (0, 7 and 14 days respectively) were separately estimated. Then we further explored the conclusion while taking the variation of *R*_0_ into account. As the vaccine efficiency and total supply may also affect the result of optimization, such two factors were also parameterized and applied in the analysis by setting the high (90%) and the low (60%) level of efficiencies and three sizes (10%, 30%, 50% of total population) of supply.

The scenario of China was primarily concerned in the main text. It was considered with a number of susceptible individuals and moderate transmission rates (Hypothetical *R*_0_ was set from 1.15 to 1.8). Chinese age demographics, the number of oversea imported travelers in the last year and existing vaccine coverage and the contact structure were all implemented at the initial step of simulation.

### Threshold conditions for herd immunity

Vaccination-induced herd immunity refers to the indirect protection for unvaccinated individuals provided by other ones with vaccine-induced immunity in a highly vaccinated population[23]. In this study, we evaluated the threshold conditions of five countries (China, America, India, South Africa and Brazil) to achieve herd immunity with vaccination when we combat the newly emerged delta variant, which hold so far the greatest transmission ability (hypothetical *R*_0_ = 5.0)[24].

Additionally, other than quarantine measures, keeping social distance and wearing masks also contributed to reducing cumulative infections and related deaths[25]. Thus such factors were implemented in order to provide valuable information for decision-making on virus elimination.

### Sensitivity analysis

Although all established assumptions were endorsed by solid evidences collected from governmental documents and highly influential literature, we mentioned that to some extent the modelling results varied with essential parameters settings, including the susceptible rate of current vaccinated population. Thus we conducted a sensitivity analysis to characterize its impact on the robustness of conclusions.

Detailed descriptions of modelling framework and parameters settings can be found in Additional file 1. All the analyses were performed by using R software(v4.0.2, R Foundation; Vienna, Austria).

## Results

### Boosting vaccine prioritization

Firstly, we introduced the mean value of infectious outliers rate under 0,7 and 14 days of quarantine respectively into the model (Table 1). For the scenario without quarantine measures, such parameter was set as 100%. By simulating the Chinese transmission (hypothetical *R*_0_ =1.15) with high vaccine efficiency (80%), we found that prioritizing high-risk workers the booster dose with at least 7-days quarantine time reaches lower cumulative infections and deaths. It also implies that when there were no restrictions implemented, prioritizing vaccinating susceptible ones was optimal as a number of normal individuals remained susceptible to infections. Besides, mass vaccination were effectively reduced the transmission severity in all scenarios of quarantine time (Fig. 2).

**Table 1:** Five essential percentiles and the mean of infectious outliers rate under four quarantine strategies.

| Quarantine strategy, days | Parameters, % of total infections |  |  |  |  |  |
| --- | --- | --- | --- | --- | --- | --- |
|  | mean | 1th percentile | 25th percentile | 50th percentile | 75th percentile | 99th percentile |
| 7 | 30.876 | 29.258 | 30.342 | 30.886 | 31.382 | 32.807 |
| 14 | 10.529 | 9.506 | 10.219 | 10.509 | 10.825 | 11.524 |
| 21 | 4.533 | 3.856 | 4.347 | 4.523 | 4.731 | 5.186 |
| 28 | 2.254 | 1.813 | 2.13 | 2.247 | 2.375 | 2.689 |

**Fig. 2:**
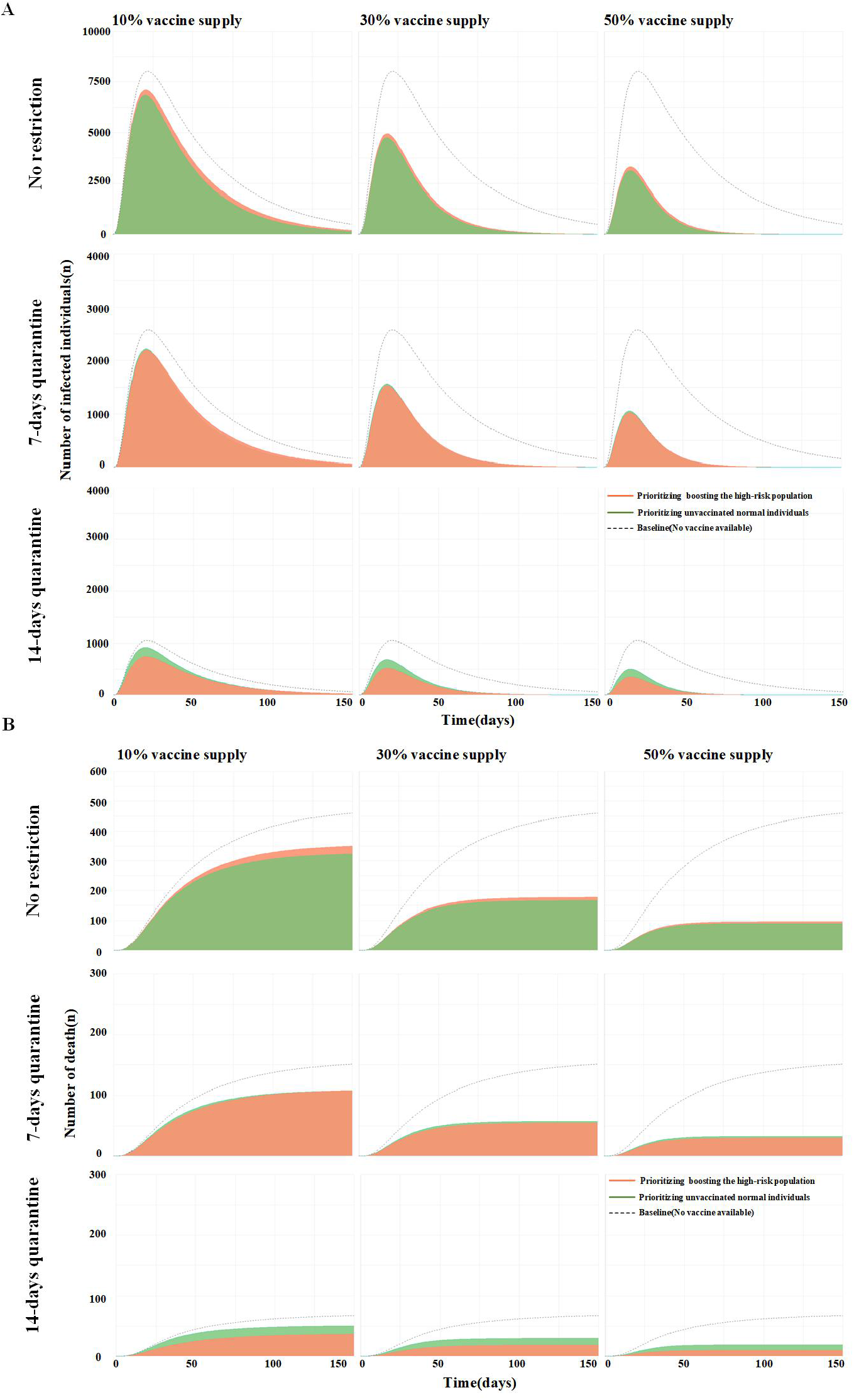
Comparison of two vaccination prioritization strategies under scenarios with different length of quarantine and vaccine supply. **A** Dynamics of hypothetical epidemic in China. In this scenario we set moderate transmission rate(basic reproduction number,*R*_0_) as 1.15, and the optimal strategy is boosting high-risk workers if at least 7-days quarantine is maintained. **B** Cumulative deaths caused by COVID-19 infection. Results showed identical conclusions in both infections and mortality simulation.

Secondly, we also explored the influence of higher transmission rate and different levels of vaccine efficiency on the optimization. Results showed that the higher transmission rate was (*R*_0_ ranged from 1.15 to 1.8), the longer quarantine time was needed to remain boosting high-risk workers as optimal strategy. Besides, when the quarantine duration was fixed, using flawed vaccine resulted in the strategy of prioritizing susceptible normal people becoming optimal (Fig. 3).

**Fig. 3:**
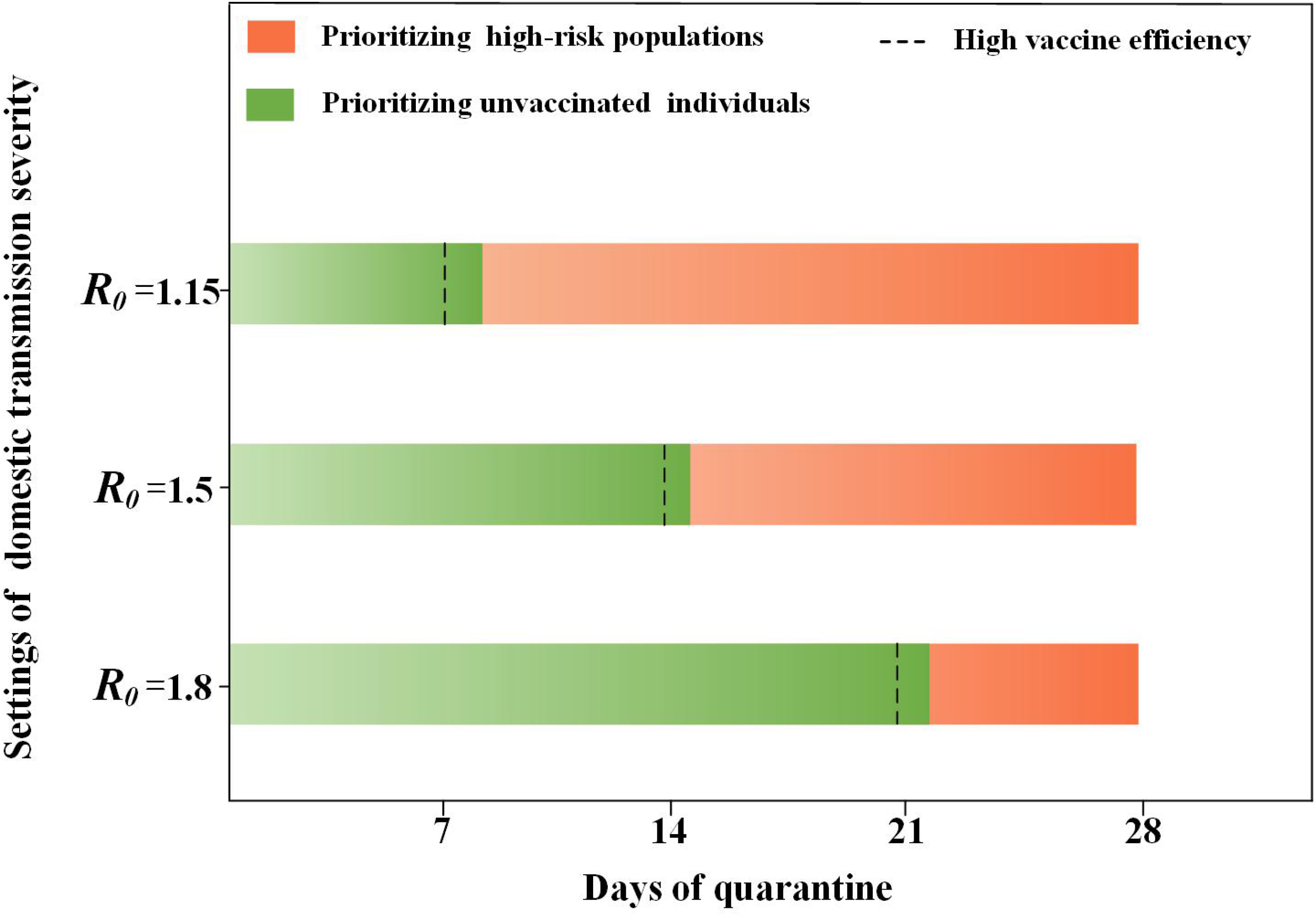
Optimal vaccination strategies under different transmission rate and quarantine settings. The dashed line represents the the least required quarantine days of taking boosting high-risk workers with high efficiency vaccine as optimal, while under the same quarantine settings, prioritizing normal susceptible individuals is optimal when flawed vaccine is applied.

Analysis above implied that the border quarantine policy not only determined the optimal domestic vaccinating strategies, but also contributed to reducing the transmission risk. However, with longer quarantine time, the corresponding reduction did not remain its upward trend throughout all transmission rate settings. We found that when the *R*_0_ lay within 1.15–1.5, the more restrictive measures correspond to the higher extent of reducing infections, and the greatest improvement between strategies was from 0 to 7-days of quarantine. By contrast, such reduction of all quarantine strategies dramatically fell afterwards and reached its minimum effect when *R*_0_ = 2.5 (Fig.4A).

**Fig. 4:**
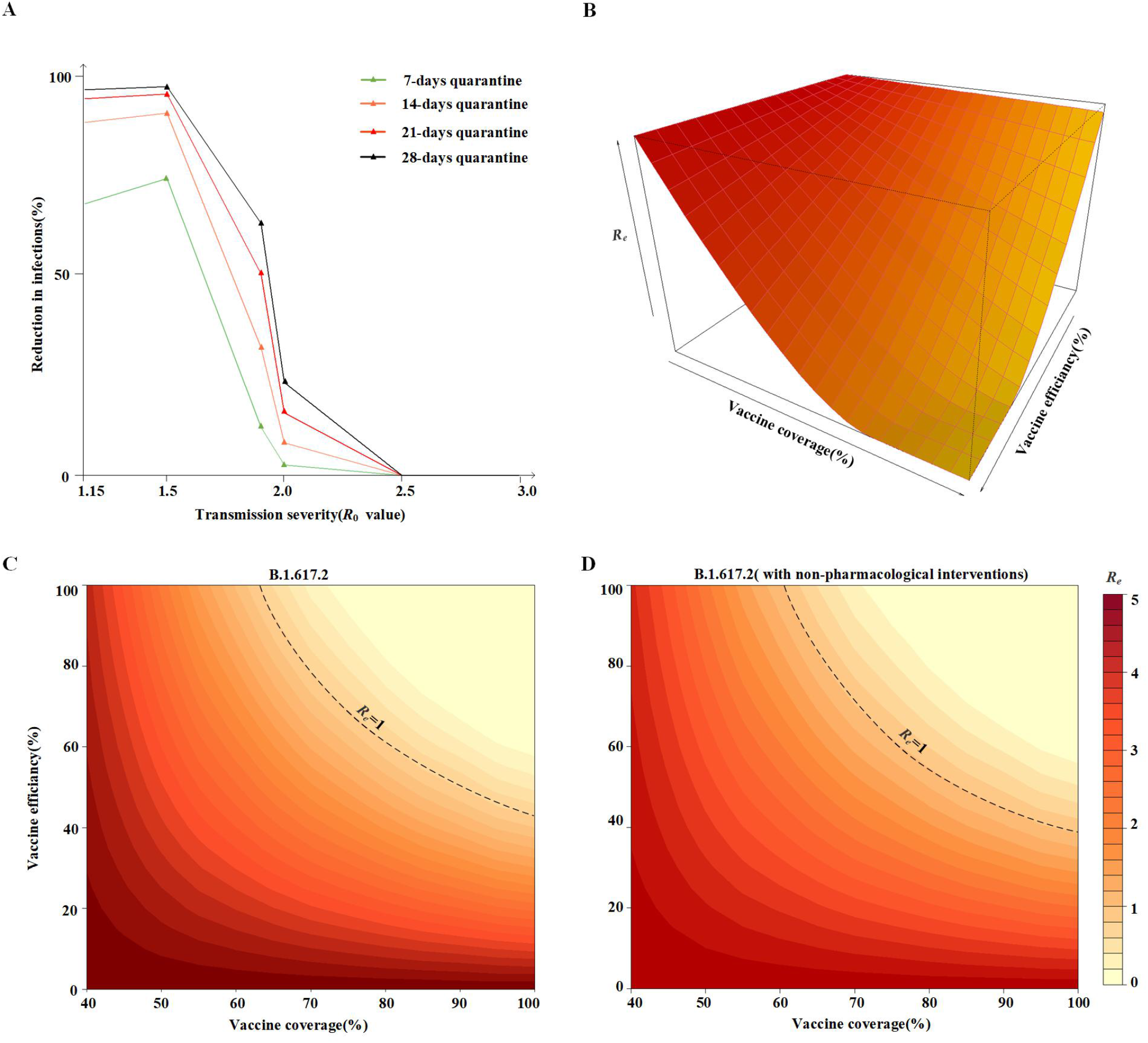
Simulation results for Chinese scenarios. **A** Reductions in infections of four quarantine durations. Without the vaccination project, maintaining one week surveillance lead to 69.12% reduction in cases compared to no restrictions. And the suggested duration should not exceed 21 days because its improvement to the 28-days is lower at 2.28%. **B** Visualizing the decrease trend of *R*_*e*_ with the growth of vaccine coverage and efficiencies. The extent of decrease mainly depends on the increase of total vaccine coverage. **C** The heat map with contours of different levels of transmission severity under varying vaccine coverage and efficiency settings. **D** The scenario considered with interventions of wearing masks and keeping social distance.

Such model-informed conclusions indicated that although it is still effective in containing transmission with mere quarantine policies in the certain range of *R*_0_, such measure have limited contribution to achieving herd immunity in confronting the highly contagious delta variant since there were always infectious outliers, and even one of them can rapidly cause a comprehensive spread. Thus in the following analysis we merely took interventions of keeping social distance and wearing masks into account.

### Threshold conditions for herd immunity

As the classical herd immunity level was oversimplified as 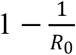[26], to better model the complex phenomenon in China, we here defined such concept as the condition leading the effective reproduction number *R*_*e*_ less than 1. By parameterizing the vaccine efficiency from 0 to 100% at the pace of 5%, we estimated the least required vaccine coverage of total population to reach the goal of herd immunity when combat the delta variant. We found that with 50% vaccine efficiency, the required coverage of vaccination was 89%, while the result is more achievable that vaccinating 74% of total people with 70% efficiency against infection can also reach the goal. Besides, the factor of vaccine coverage influenced more than its efficiency on the transmission reduction (Fig.4BC).

According to the evidence that keeping social distance as well as wearing masks can provide at least 24.3% joint reduction in infections[25], we further explored their impact on accelerating the process of approaching herd immunity. Results showed that under the same vaccine efficiency settings, applying such measures can effectively lower the threshold requirement of vaccine coverage to accomplish herd immunity. To be specific, vaccinating 84% and 71% (corresponding to 50% and 70% vaccine efficiency respectively) were sufficient to reach the goal if interventions are implemented (Fig.4D).

We also estimated the situation of four other countries, which were considered with different age demographics, contact structures, existing vaccination coverage and seroprevalence. Results of other countries were shown in Table 2 and Fig. 5. In addition, to more conservatively estimate the threshold conditions to herd immunity, we performed the sensitive analyses by applying a higher rate (60%) of existing susceptible individuals among vaccinated ones. Such results did not change significantly and detailed information can be found in Additional file 1.

**Table 2:**
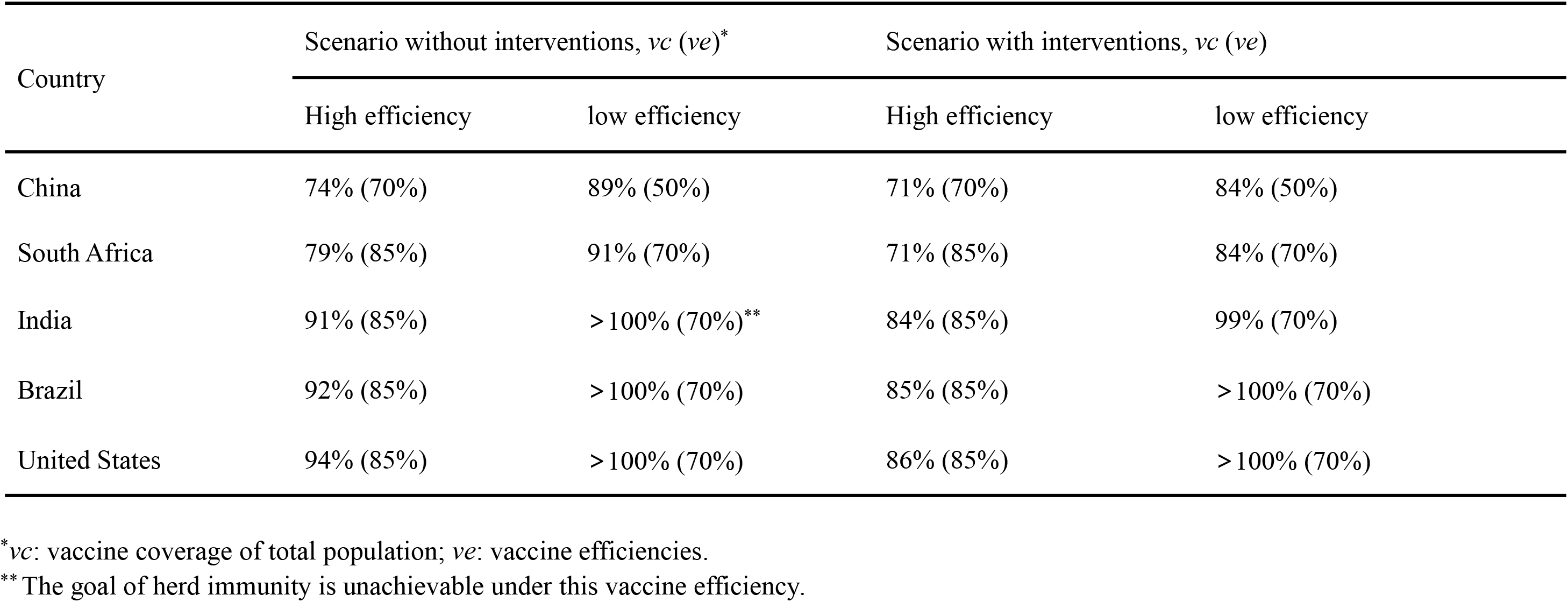
Threshold vaccine coverage of five countries to reach herd immunity with varying vaccine efficiencies.

**Fig. 5:**
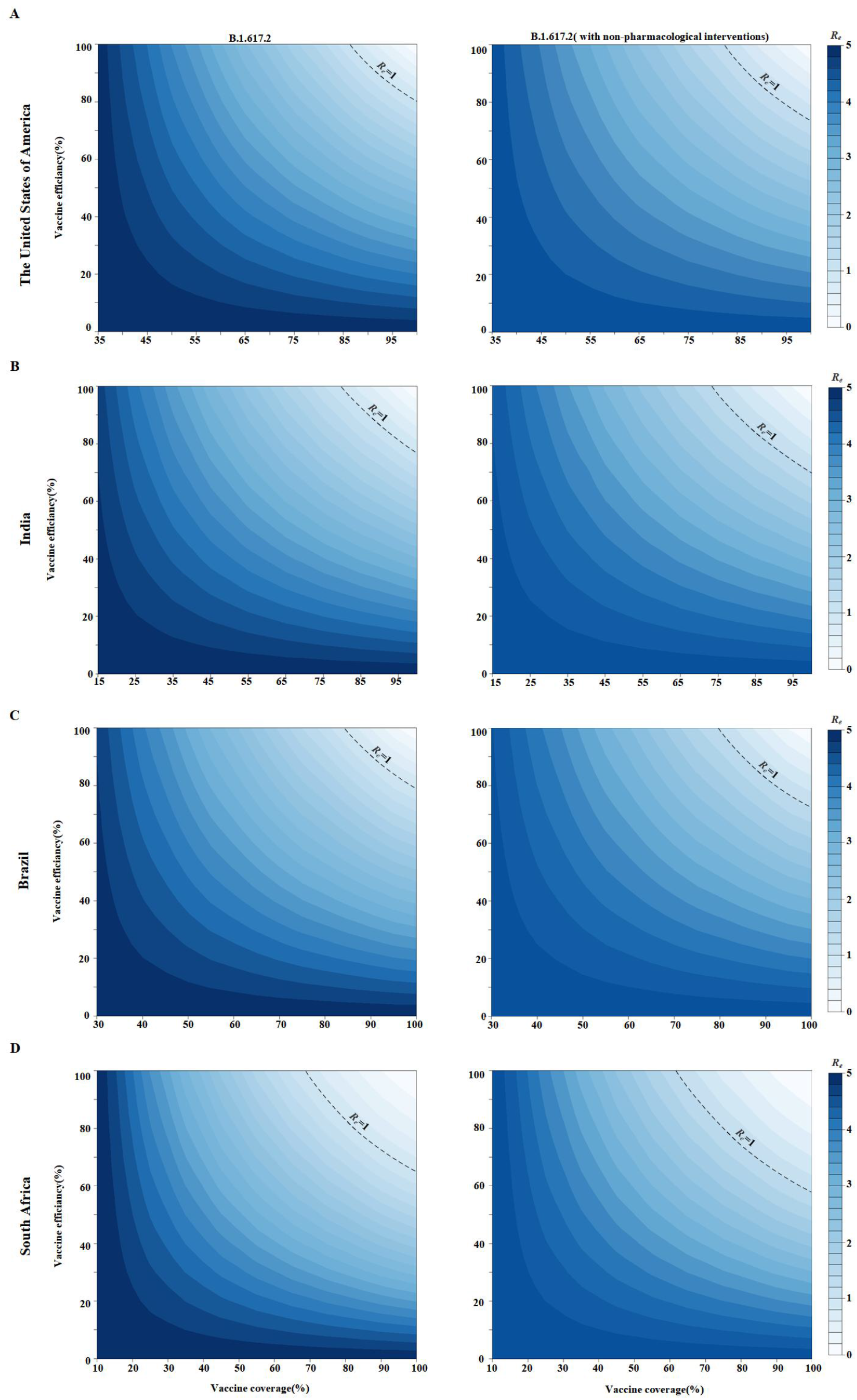
Simulation results for other countries. We also considered the seroprevalence for such countries due to their previous community COVID-19 spread. Such individuals with naturally acquired or vaccine-induced immunity were assumed that 50% of them remained perfect protection against infection and the rest was considered as susceptible ones. **A** United States scenarios. **B** India scenarios. **C** Brazil scenarios. **D** South Africa scenarios.

## Discussion

Two major subjects we account for in this study were: (1) informing decision-makers that distributing vaccine to which groups first, high-risk individuals or susceptible normal ones was optimal to reach the lower COVID-19 transmission risk and (2) The specific vaccine coverage that can lead us to the promising herd immunity. By using the proposed novel model, we unprecedentedly addressed such problem while also took the complex effect of different non-pharmacological interventions into account. According to our modelling results, at least 7-days quarantine time should be maintained so that boosting high-risk workers was more effective in reducing the total infections and deaths. Besides, vaccinating 74%–89% of total populations with varying efficiency (70%–50%) can help accomplish the virus-eliminating project of China, which was generally in consistent with the figure suggested by World Health Organization (WHO) and previous estimation conducted by Liu et al[27,28]. Additionally, the required coverage decreased to 71%–84% when the restrictions of keeping social distance and masks wearing were implemented. In fact, unlike keeping social distance, wearing masks was relatively more practicable and less harmful to the economics. Thus we solely incorporated the parameter of mask use into the model, and the result was slightly lower at 72%–86%. Such evidences were also valuable for countries in similar situations.

In this study we also creatively proposed the infectious outliers distribution, and their rate under different quarantine durations can thus be quantified. Our results indicated that a longer time of screening can effectively reduced more transmission risk when the COVID-19 was moderately prevalent. As the improvement between strategies was becoming minimum, we suggested the screening policy could be maintained bellow 21 days according to the cost-effectiveness.

Although by merely using the quarantine policy we were unable to diminish COVID-19 when the transmission rate was high, since the mass vaccination significantly alleviated the transmission severity, restrictive quarantine measures combined with comprehensive vaccine-induced protection enabled us to reduce imported infections to the minimum level. Our findings well supported the current practice of maintaining the restrictions on international travel in China, which hold the belief that health is above wealth[29]. On the other hand, we wondered if there was one individualized quarantine method, which can not only reduce infections but also dramatically shorten the overall surveillance time due to a number of people identified as low-infection risk so that they will be released earlier, thus the process of reopening the external gate of global countries will be highly accelerated. Such idea is achievable once we provide the tool of qualifying the quarantine efficiency.

A number of published studies focused on the subject of vaccine prioritization[30,31,32]. Bubar et al.[19] considered such optimization problem with the existing serostatus of people in United States, and provided useful tools to simulating COVID-19 transmission with numerous parameters that might affect the result. However, to the best of our knowledge, handful studies examined the current practice of boosting specific populations aimed to acquire the further reduction of transmission risk and infections. It has been taken as a common sense that compared to normal people, those who exposed the most due to the occupation, as well as the most susceptible ones with worse consequences once being infected, should be protect first with vaccination[2]. Since a large number of high-risk workers have been fully vaccinated, according to the result of our study, the risk posed by existing infections on normal susceptible people will higher than that on high-risk workers as long as the susceptible individuals are widely distributed and there are no quarantine measures maintained, which means under such scenarios, prioritizing the former is optimal to reach minimum transmission.

Chinese health administration previously announced that 1.04 billion people in china had been fully vaccinated, which meant the vaccine coverage already reached the threshold line of herd immunity[33]. However, due to imperfect vaccine efficiency against infections as well as decreasing vaccine-induced protection over time, the threshold condition proposed in this study should be interpreted as the current proportion of people with sufficient protection. Thus we suggest that the following surveillance should be implemented to investigate the prevalence of antibody levels among vaccinated population, so that we can ensure the real-time number of susceptible individuals. Additionally, as United States hold the highest required vaccine coverage (94%) to reach herd immunity and the largest volume of international entrants (86.1 million)[34], we wonder that once we remove the border restrictions, whether the increasing number of such individuals will lead to the higher requirement of vaccination proportion. Thus the number of imported travelers in the year before COVID-19 pandemic was applied to simulate the scenario of reopening the national border. Fortunately, results did not change significantly and proved the robustness of our estimation.

Several limitations of the present study exist. Firstly, we merely evaluated the optimal strategy between prioritizing the high-risk workers or the susceptible normal individuals, which can be more specific if the latter option was preferred. Following research can further explore the best vaccination strategy among different age groups by using our proposed model. For example, evidences showed that prioritizing the 20–49 age group and adults greater than 65 years old can reach the least infections and deaths respectively[19]. Considering with different interventions and the various existing vaccine coverage, the question that whether such conclusions are still valid is yet to be answered. Secondly, conclusions in this study are simply interpretation of simulation results. Such model-informed instructions need to be further verified by real-world research.

## Conclusions

Non-pharmacological interventions can substantially affect booster vaccination prioritization and the threshold condition of vaccine coverage to reach herd immunity. First boosting high-risk workers is optimal to minimize infections and deaths in China if at least 7-days of quarantine is maintained. As the improvement between quarantine strategies was becoming insignificant, current suggested duration is bellow 21 days due to the cost-effectiveness. Besides, mass vaccination combined with restrictive quarantine measures can help reduce the transmission risk caused by imported infections to the minimum level.

On the other hand, despite the varying country-specific requirement of vaccination proportion to reach herd immunity, wearing masks and keeping social distance can effectively lower such threshold condition to accomplish the goal of herd immunity. However, due to the imperfect vaccine efficiency, as well as the vaccine-induced protection that decreased with time, the following surveillance of vaccinated populations is necessary to ensure the real-time proportion of vaccinated individuals with sufficient protection and the number of susceptible ones who need the booster dose.

## Data Availability

All data and parameters used are publicly available.

## Data Availability

All data and parameters used are publicly available.

## Authors’ contributions

Model design and methodology: ZY L, T W; Simulation: ZY L, BR Y, JL W; Parameters and data collection: BR Y, JL W, WJ L; writing and drawing: ZY L, BR Y, JL W. All authors have read and approved the final manuscript.

## Acknowledgments

We would like to thank the health commission of Shanxi province for the grant of major science and technology project of Shanxi province (Grant number: 202005D121008) of Shanxi department of science and technology and the educational innovation project of Shanxi province(No.40). We also appreciate the code consultation provided by Jinsong Wu from Xi’an University of Posts and Telecommunications.

## Ethics approval and consent to participate

Not applicable

## Consent for publication

Not applicable

## Supplementary materials

We offered our extended information mentioned in the main text and related references in the Additional file 1.

