## Additional file 1 for "Impact of non-pharmacological interventions on COVID-19 boosting vaccine prioritization and vaccine-induced herd immunity: a population-stratified modelling study"

### Supplementary Appendix

#### Definition of infections and high-risk population

**Types of Infection**

Case definitions were based on the Protocol for COVID-19 Prevention and Control of the National Health Commission of China[1].

1. Symptomatic infections: patients who tested positive by RT－PCR and showed symptoms before presenting to a hospital during the 2-week quarantine period as a close contact, during the hospital stay or within 4 weeks after being discharged from the hospital. An RT－PCR cycle threshold value (Ct value) of less than 37 was defined as positive, using a commercial RT－PCR kit (DAAN Gene, 20203400063).

2. Asymptomatic infections: patients who tested positive by RT－PCR and reported no symptoms before being diagnosed and did not show symptoms throughout the quarantine and treatment period and in 4 weeks after being discharged from the hospital. Of note, cases without symptoms upon diagnosis were labeled as ‘asymptomatic’ temporarily; this label was revised to ‘symptomatic’ later if the cases developed symptoms.

**High-risk population related to the closed-loop management**

Occupation with high risk of exposure to COVID-19 infections are listed bellow[2]:

1. Aviation faculties: Including individuals working or living in airplane or airport.
2. Health care workers: including doctors, nurses, diseases control related workers, health department faculty and public health organizations.
3. Community workers: providing essential support for quarantined individuals.
4. Security personnel: including police and security team.
5. Staff at quarantine facilities: including receptionists and other hotel service personnel.

#### Characteristics of patients applied to estimate infectious outliers distribution

**Table S1: Sex, age demographics and location characteristics of total 1920 cases.**

| Characteristics | All (*n* = 1920) | Symptomatic (*n* = 1745) | Uncommon infections | | | |
|---|---|---|---|---|---|---|
| | | | Presymptomatic (*n* = 20) | Asymptomatic (*n* = 105) | Recurrent positive (*n* = 50) | Total (*n* = 175) |
| Sex, n (%) | | | | | | |
| Male | 933 (48.6) | 86 (49.3) | 1 (5.0) | 36 (34.3) | 36 (72.0) | 73 (41.7) |
| Female | 707 (36.8) | 664 (38.1) | 1 (5.0) | 31 (29.5) | 11 (22.0) | 43 (24.6) |
| unknown | 280 (14.6) | 221 (12.6) | 18 (90.0) | 38 (36.2) | 3 (6.0) | 59 (33.7) |
| Age groups, n (%) | | | | | | |
| 0-14 | 53 (2.8) | 39 (2.2) | 0 (0.0) | 9 (8.6) | 5 (10.0) | 14 (8.0) |
| 15-64 | 1389 (72.3) | 1302 (74.6) | 2 (10.0) | 48 (45.7) | 37 (74.0) | 87 (49.7) |
| ⩾ 65 | 124 (6.5) | 118 (6.8) | 0 (0.0) | 1 (1.0) | 5 (10.0) | 6 (3.4) |
| unknown | 354 (18.4) | 286 (16.4) | 18 (90.0) | 47 (44.7) | 3 (6.0) | 68 (38.9) |
| Location, n (%) | | | | | | |
| The mainland of China | 1557 (81.1) | 1413 (81.0) | 19 (95.0) | 75 (71.4) | 50 (100.00) | 144 (82.3) |
| Outside the mainland of China | 118 (6.1) | 105 (6.0) | 1 (5.0) | 12 (11.4) | 0 (0.00) | 13 (7.4) |
| Unknown | 245 (12.8) | 227 (13.0) | 0 (0.0) | 18 (17.2) | 0 (0.00) | 18 (10.3) |

#### Model design

A continuous-time ordinary differential equations (ODE) compartmental model stratified by populations and age was used to quantify the impact of non-pharmacological interventions on boosting vaccine prioritization and herd immunity. As figure 1 in the main text illustrate the modeling framework, major ordinary differential equations are as follow:

$$\frac{dE_o}{dt} = (1-r)S_o - \omega E_o$$

$$\frac{dS_h}{dt} =- (1 - ve \times vc)\lambda_h S_h$$

$$\frac{dS_i}{dt} =- (1 - ve \times vc)\lambda_i S_i$$

Where the $S_o$ , $S_h$ , $S_i$ are the total number of travelers, high-risk workers and normal individuals of China respectively, and $r$ represent the infections rate among passengers. $ve \times vc$ is the proportion of individuals who acquired perfect protection once they receive the shot according to the all-or-nothing assumption listed in the main text, where $ve$ is the vaccine efficiency and $vc$ is the vaccine coverage of total supply.

The infections force for an individual in high-risk populations is $\lambda_h = u_h\left(c_{ho}\frac{I_o}{N_o-\Omega_o} + c_{hh}\frac{I_h}{N_h-\Omega_h}\right)$ where the $u_h$ is the susceptibility that successfully being infected after contact with an infections, and $c_{ho}$ and $c_{hh}$ are the contact number per day of high-risk populations with travelers and itself respectively. The following term $\frac{I_o}{N_o-\Omega_o}$ and $\frac{I_h}{N_h-\Omega_h}$ represent the current proportion of infections among all living individuals of travelers and high-risk populations, and the $I_o$, $N_o$, $\Omega_o$ and $I_h$, $N_h$, $\Omega_h$ are the number of infections, total number of individuals and deaths of such two populations respectively.

The parameter $\lambda_i = u_i\left(c_{io}\frac{I_o}{N_o-\Omega_o} + c_{ih}\frac{I_h}{N_h-\Omega_h} + \sum_j c_{ij}\frac{j}{N_j-\Omega_j}\right)$ represent the infections force for individuals in normal population, where the $c_{io}$, $c_{ih}$, and $c_{ij}$ respectively represent the contacts with travelers, high-risk workers and individuals in other age groups( $i = 1,2,3, ... 9$) among normal populations.

To incorporate the impact of border quarantine policy, the infectious outliers rate (also can be interpreted as quarantine efficiency, $qe$) under different lengths of such measures were calculated and implemented into the model. Then the force of infection for normal population is:

$$\lambda_i = u_i \left( c_{io} \frac{I_q}{N_o - \Omega_o} + c_{ih} \frac{I_h}{N_h - \Omega_h} + \sum_j c_{ij} \frac{j}{N_j - \Omega_j} \right)$$

Where $I_q = I_o * qe$ is the number of travelers who remain infectious after the specific quarantine duration.

In this study we define herd immunity as the condition that make the effective production number $R_e$ reached 1. We first calculate the maximum infection number of the simulation when combat the delta variant without vaccination. It was considered as the baseline scenario corresponding to $R_0$=5, and we then take the vaccination of varying efficiencies and coverage into account in order to quantify its impact on reduction in transmission rate.

$$R_e = \frac{max[sum(I_v)]}{max[sum(I_b)]} \times R_0$$

Where $sum(I_v)$ and $sum(I_b)$ were the sum of infections with and without vaccination project respectively. With the 24.3% joint reduction in infection of interventions including wearing masks and keeping social distance, then the $R_e$ equation is：

$$R_e = \frac{max[sum(I_c)]}{max[sum(I_b)]} \times R_0$$

Where $I_c = I_v * 0.757$ is the number of infections with combined effect of interventions and vaccination.

Although the corresponding infection number was not mathematically equal to zero when $R_e$ reached the level of herd immunity, it can be interpreted that the transmission definitely remain the downward trend and due to the significant delays between infections and interventions the number of incidence will not reach zero immediately.

#### Supplemental assumptions

**Vaccination:**

(A1).The delay between vaccination and protection is precede the simulation;

(A2).The existing vaccine coverage were assumed mostly distributed to individuals aged in 20-70(See specific settings in section 6, page 15);

(A3).The high-risk workers were already fully vaccinated and were considered with lower susceptibility, the follow vaccination was the booster dose(See specific settings in section 6, page 15);

**Contact matrix:**

(A5).The original contact matrix characterized the situations without non-pharmacological interventions and before the transmission;

**Table S2: The structure of contact matrix**

| | 0-9 | 10-19 | 20-29 | 30-39 | 40-49 | 50-59 | 60-69 | 70-79 | 80+ | HRW* | T** |
|---|---|---|---|---|---|---|---|---|---|---|---|
| 0-9 | $c_{11}$ | $c_{12}$ | $c_{13}$ | $c_{14}$ | $c_{15}$ | $c_{16}$ | $c_{17}$ | $c_{18}$ | $c_{19}$ | $c_{1h}$ | $c_{1o}$ |
| 10-19 | $c_{21}$ | $c_{22}$ | $c_{23}$ | $c_{24}$ | $c_{25}$ | $c_{26}$ | $c_{27}$ | $c_{28}$ | $c_{29}$ | $c_{2h}$ | $c_{2o}$ |
| 20-29 | $c_{31}$ | $c_{32}$ | $c_{33}$ | $c_{34}$ | $c_{35}$ | $c_{36}$ | $c_{37}$ | $c_{38}$ | $c_{39}$ | $c_{3h}$ | $c_{3o}$ |
| 30-39 | $c_{41}$ | $c_{42}$ | $c_{43}$ | $c_{44}$ | $c_{45}$ | $c_{46}$ | $c_{47}$ | $c_{48}$ | $c_{49}$ | $c_{4h}$ | $c_{4o}$ |
| 40-49 | $c_{51}$ | $c_{52}$ | $c_{53}$ | $c_{54}$ | $c_{55}$ | $c_{56}$ | $c_{57}$ | $c_{58}$ | $c_{59}$ | $c_{5h}$ | $c_{5o}$ |
| 50-59 | $c_{61}$ | $c_{62}$ | $c_{63}$ | $c_{64}$ | $c_{65}$ | $c_{66}$ | $c_{67}$ | $c_{68}$ | $c_{69}$ | $c_{6h}$ | $c_{6o}$ |
| 60-69 | $c_{71}$ | $c_{72}$ | $c_{73}$ | $c_{74}$ | $c_{75}$ | $c_{76}$ | $c_{77}$ | $c_{78}$ | $c_{79}$ | $c_{7h}$ | $c_{7o}$ |
| 70-79 | $c_{81}$ | $c_{82}$ | $c_{83}$ | $c_{84}$ | $c_{85}$ | $c_{86}$ | $c_{87}$ | $c_{88}$ | $c_{89}$ | $c_{8h}$ | $c_{8o}$ |
| 80+ | $c_{91}$ | $c_{92}$ | $c_{93}$ | $c_{94}$ | $c_{95}$ | $c_{96}$ | $c_{97}$ | $c_{98}$ | $c_{99}$ | $c_{9\mathrm{h}}$ | $c_{9o}$ |
| HRW | $c_{h1}$ | $c_{h2}$ | $c_{h3}$ | $c_{h4}$ | $c_{h5}$ | $c_{h6}$ | $c_{h7}$ | $c_{h8}$ | $c_{h9}$ | $c_{hh}$ | $c_{ho}$ |
| T | $c_{o1}$ | $c_{o2}$ | $c_{o3}$ | $c_{o4}$ | $c_{o5}$ | $c_{o6}$ | $c_{o7}$ | $c_{o8}$ | $c_{o9}$ | $c_{oh}$ | $c_{oo}$ |

*HRW: high-risk workers

**T: travelers

(A6).The contact rate of travelers with nine age groups of the normal population, high-risk workers and itself was set as zero;

$$c_{oi} = c_{oh} = c_{oo} = 0$$

Because the derivatives of susceptible compartment is fixed with infectious rate $r$ corresponding to the closed-loop management that the travelers population can not be infected by high-risk workers, normal population and by each other.

(A7).The contact rate of nine age groups with high-risk workers was assumed as their contact frequencies with the specific age group which hold similar proportion of high-risk workers;

$$c_{ih} = c_{ij}$$

Where the age group-j hold similar proportion with high-risk workers.

(A8).The contact rate of nine age groups with travelers was assumed as the mean value of the number of each age group with itself other eight ones;

$$c_{io} = \frac{\sum_j c_{ij}}{9}$$

Corresponding to the fact that if no quarantine measures and transmission were implemented then the contact between them is as usual.

(A9).The contact rate of high-risk workers with nine age groups, itself and travelers were assumed as zero, the sum of the number of nine age groups and the maximal number of nine age groups respectively;

$$c_{hi} = 0$$

$$c_{hh} = \sum_i c_{io}$$

$$c_{ho} = max(c_{io})$$

We assumed that such high-risk workers will not be infected by normal individuals and due to the occupation they contact more frequently each other. The last equation corresponds to the first assumption listed in the main text.

(A10).If quarantine polices were implemented, the specific outliers rate under different strategies, which refer to the quarantine efficiency parameter $qe$, were applied to multiply the contact rate of nine age groups. On the other hand, we assumed the current contact number of high-risk worker is five-fold of the original one when the 7-days quarantine was implemented, and the following fold of contact frequencies is equal to that of quarantine days.

$$c_{q(io)} = qe \times c_{io}$$

$$c_{q(ho)} = 5x \times c_{ho}$$

Where $x = 1, 2, 3, 4$ (corresponding to 7, 14, 21, 28-days of quarantine respectively). The second equation represented the dramatic increase of the contact of high-risk workers with travelers if the longer quarantine time was implemented.

**Transmission:**

(A11). For countries other than China, we considered them with ongoing community spreading thus their fundamental $R_0$ calculated by using next generation method were assumed relatively higher;

(A12). For countries other than China, individuals who once being infected with naturally acquired immunity were assumed that 50% of them remained sufficient immunity against the reinfection(See specific settings in section 6, page 15).

#### Sensitivity analysis

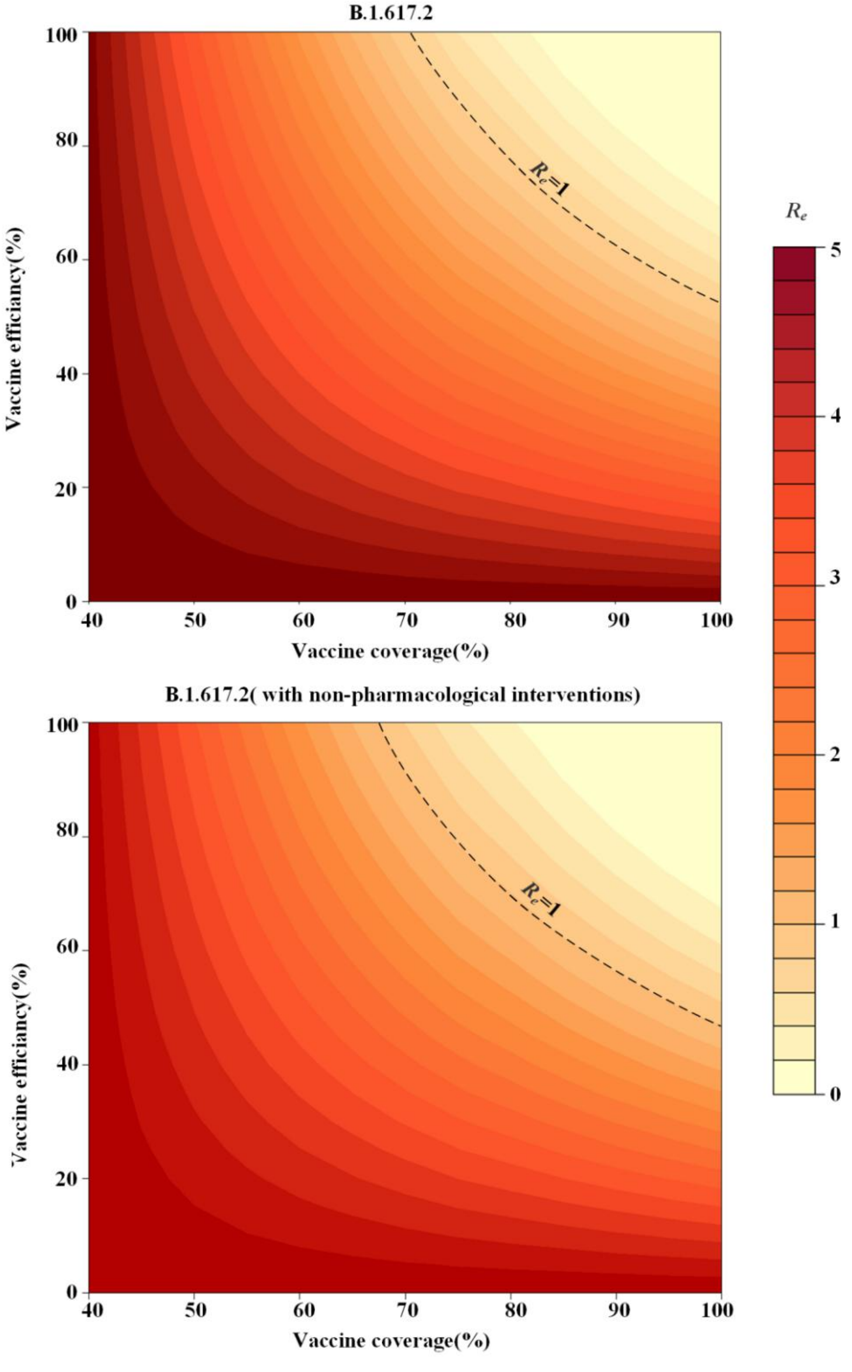

**Fig. S1: Sensitive analyses for Chinese scenarios.** In this part we applied the higher hypothetical proportion(60%) of individuals who already vaccinated but turn to susceptible due to decreasing protection over time. Results showed slightly higher conditions to reach herd immunity. Aside from the efficiency against infections, such results indicated that to take human out of the loop of boosting vaccination, we should also improve the lasting protection of vaccine.

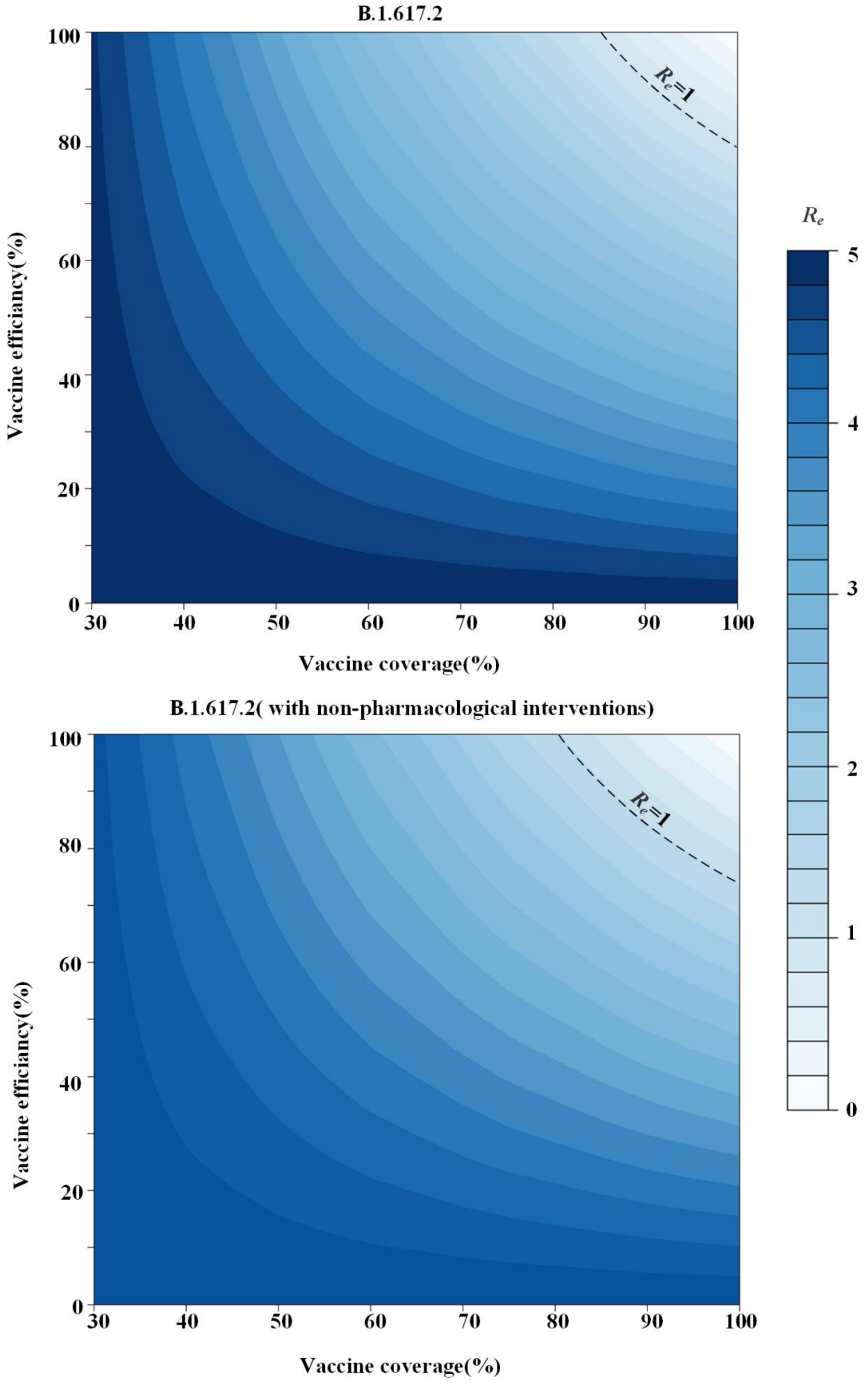

**Fig. S2: Sensitive analyses for United States scenarios.**

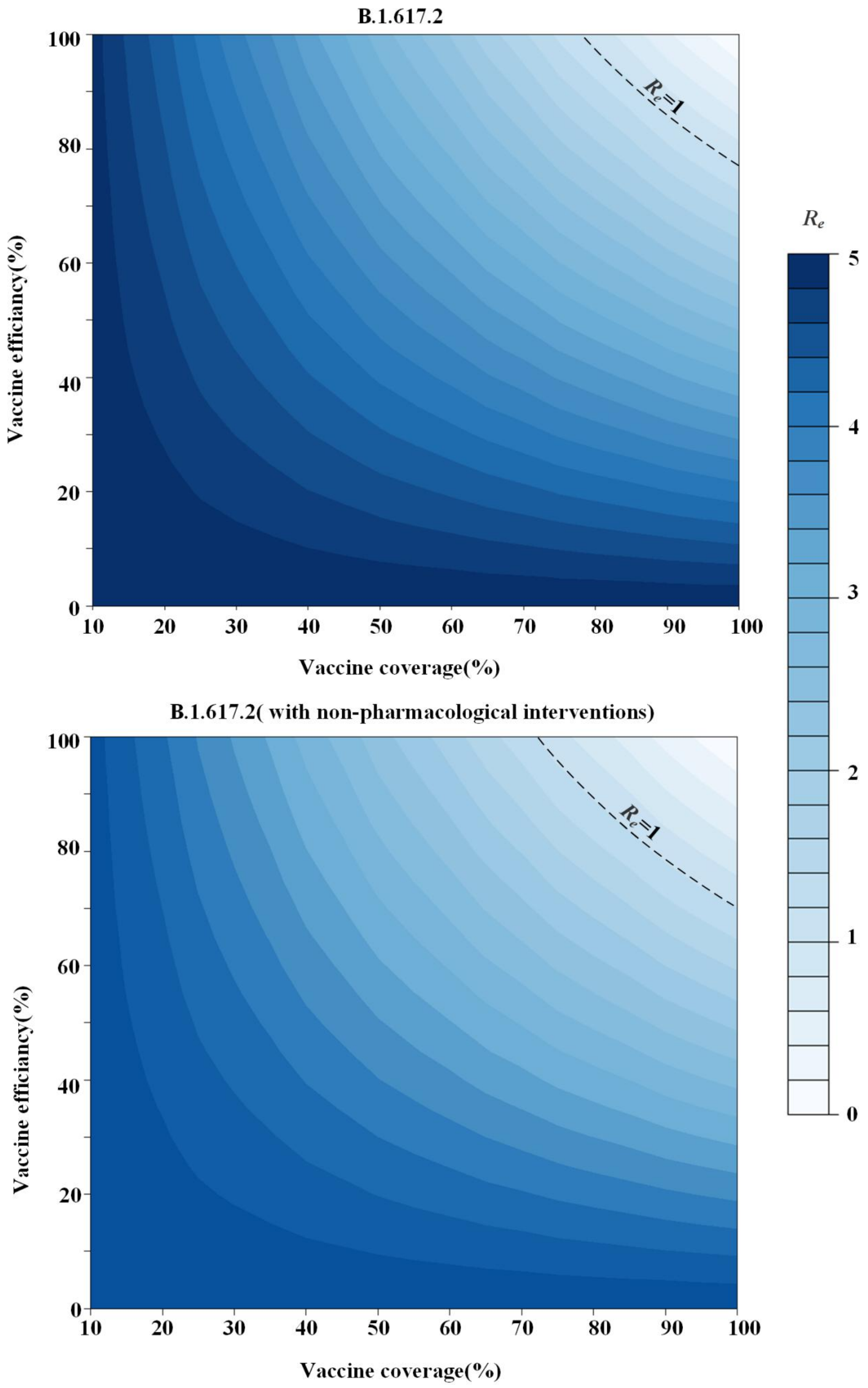

**Fig. S3: Sensitive analyses for India scenarios.**

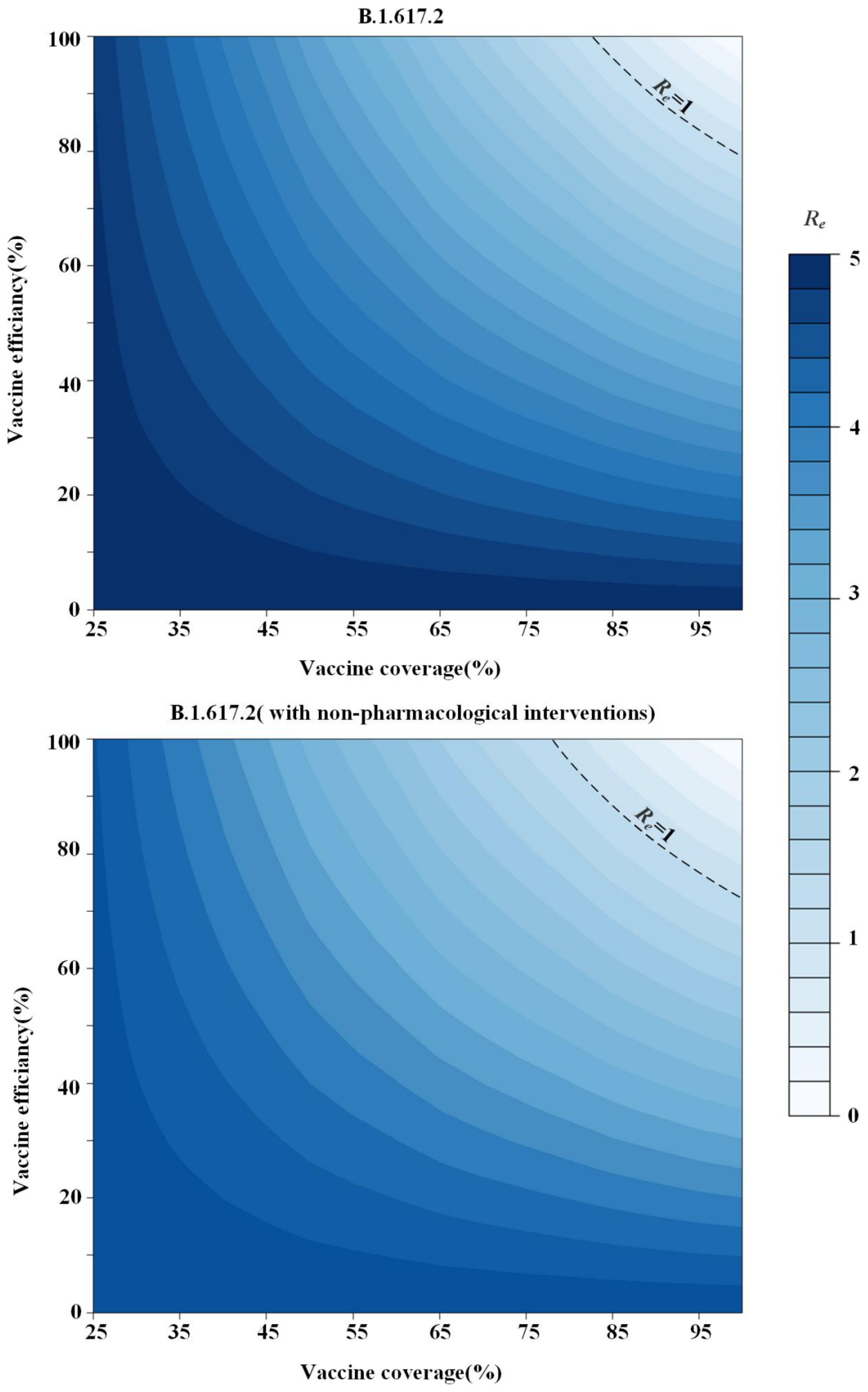

**Fig. S4: Sensitive analyses for Brazil scenarios.**

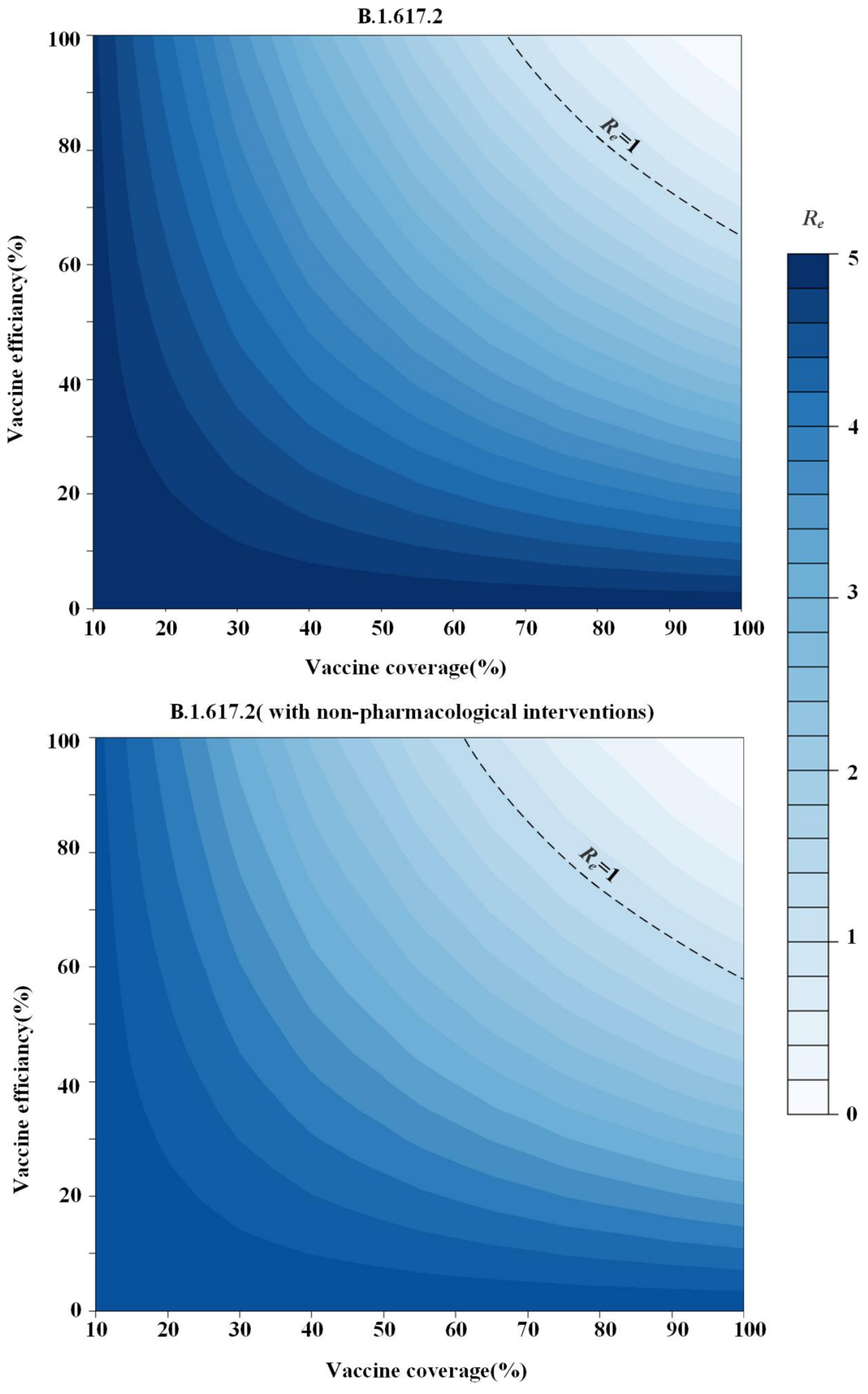

**Fig. S5: Sensitive analyses for South Africa scenarios.**

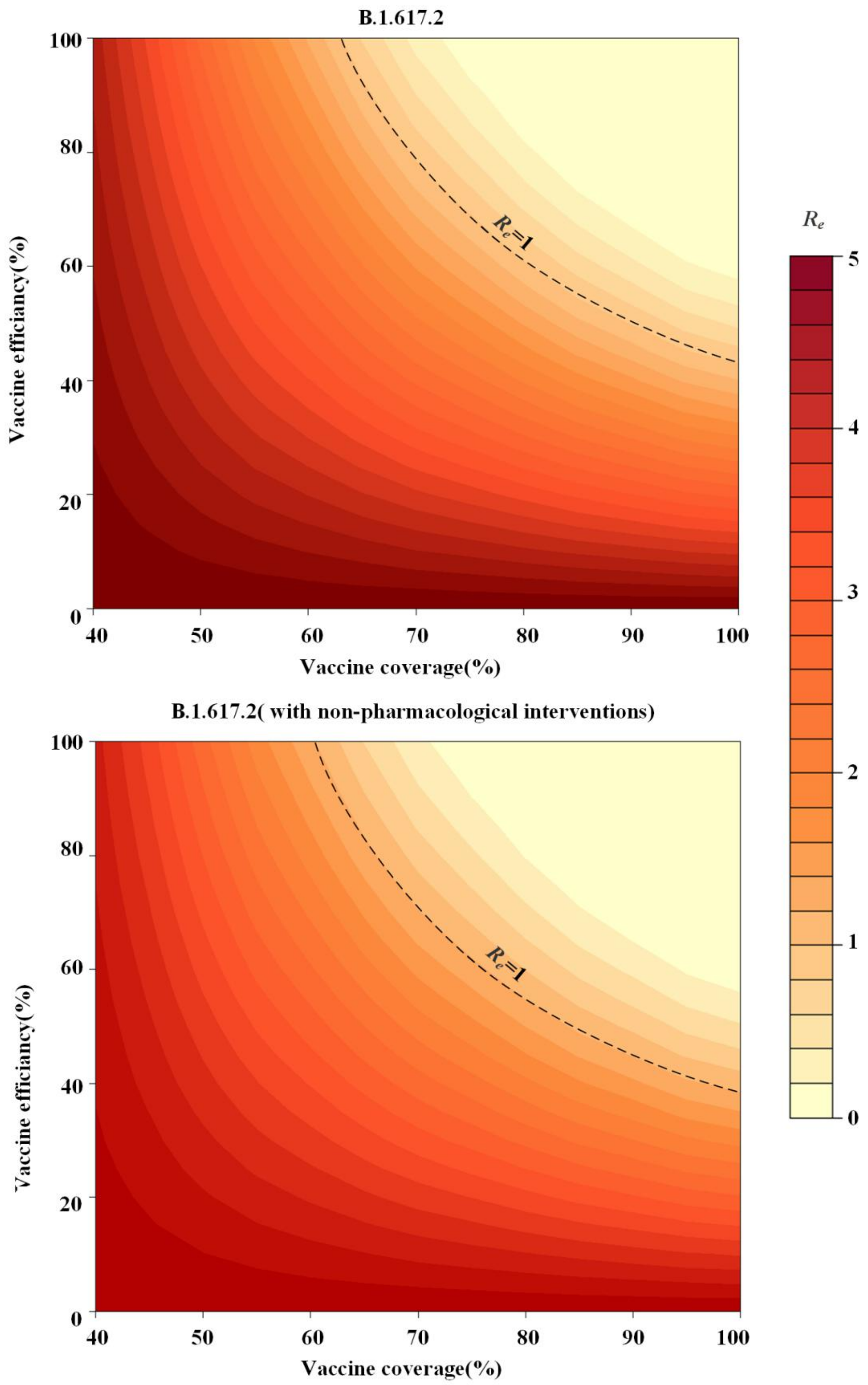

**Fig. S6: Estimations of pre-pandemic Chinese scenarios.** The number of Chinese imported travelers in the year before the COVID-19 pandemic was applied to examine the robustness of our estimation that whether the required vaccine coverage to herd immunity is varying with such number. Results showed that it is generally in consistent with the estimates in the main text.

#### Parameters settings and sources

**Table S3:Summary of parameters used in modeling and simulation.**

| Parameter | Description | Value | Source |
|---|---|---|---|
| $\omega$ | Latent and incubation period | 4.4 days | [3] |
| $\gamma$ | Infectious period | 5 days | [4] |
| $u_i$ | Susceptibility to infection for age-*i* individuals | [0.4, 0.38, 0.79, 0.86, 0.8, 0.82, 0.88, 0.74, 0.74] | [4] |
| $u_h$ | Susceptibility to infection for high-risk workers | 0.08 | [5] |
| *IFR* | Infection fatality rate | [0.001, 0.003, 0.01, 0.04, 0.12, 0.40, 1.36, 4.55, 15.24] | [4] |
| $IFR_h$ | Infection fatality rate of high-risk workers | 0.019 | [6] |
| $P_i$ | Proportion of people in age group *i* | country-specific demographic data | Table S3 |
| $P_h$ | Proportion of high-risk workers | country-specific data | Table S3 |
| $N$ | Number of total population | country-specific data | Table S3 |
| $N_I$ | Number of imported travelers | country-specific data | Table S3 |
| *snm* | Effect of keeping social distance and wearing mask | 0.757 | [7] |
| *ec* | Existing vaccine coverage | country-specific vaccination data | Table S3 |
| *r* | Infections rate of importations | 0.00016 | [8] |
| *sp* | Existing seroprevalence | country-specific | Table S3 |

| | | | |
|---|---|---|---|
| | | vaccination data | |
| $c_{ij}$ | Number of age-*j* individuals contacted by an age-*i* individual per day | country-specific contact matrix | [4] |
| $ep^*$ | Individuals with sufficient protection | $(ec+sp)*0.5$ | (A2),(A12) |

*Based on the third major assumption and supplemental ones (A2) and (A12), we calculated the country-specific distribution of *ep*.

**Table S4 : Summary of country-specific data.**

| **Country** | **parameter** | **value** | **Source** |
|---|---|---|---|
| **China** | $P_i$ | [0.11857486, 0.11575200, 0.12863483, 0.15898468, 0.15014812, 0.15436824, 0.10537164, 0.04967215, 0.01849348] | [4] |
| | $P_h$ | 0.0157 | [9] |
| | $N$ | 1443497378 | [10] |
| | $N_I$ | 27610000 | [11] |
| | $ec$ | 10478720000 | [12] |
| | $ep$ | [0.231, 0.231, 0.323, 0.323, 0.323, 0.323, 0.323, 0.323, 0.231, 0.231] | (A2),(A12) |
| **USA** | $P_i$ | [0.12000352，0.12789141，0.13216201，0.12785448，0.11480361，0.12724997，0.11627753，0.07275651，0.03971927] | [4] |
| | $P_h$ | 0.0212817 | [13] |
| | $N$ | 332915074 | [14] |

| | | | |
|---|---|---|---|
| | $N_I$ | 86100000 | [15] |
| | $ec$ | 184852416 | [16] |
| | $sp$ | 43618627 | [17] |
| | $ep$ | [0.277，0.277，0.388，0.388，0.388，0.388，0.388，0.277，0.277] | (A2),(A12) |
| **India** | $P_h$ | [0.170189049, 0.182754142, 0.172441529, 0.156485768, 0.121441764, 0.092849147, 0.063152561, 0.028387879, 0.009626253] | [4] |
| | $P_h$ | 0.0042 | [18] |
| | $N$ | 1355000000 | [19] |
| | $N_I$ | 17914000 | [20] |
| | $ec$ | 24045978 | [21] |
| | $sp$ | 33791061 | [17] |
| | $ep$ | [0.06, 0.06, 0.12, 0.12, 0.12, 0.12, 0.12, 0.06, 0.06] | (A2),(A12) |
| **South Africa** | $P_i$ | [0.195344139, 0.175508412, 0.16969035, 0.16992363, 0.117451564, 0.082813025, 0.053355436, 0.024887668, 0.007111841] | [4] |
| | $P_h$ | 0.003913927 | [22] |
| | $N$ | 60142978 | [23] |
| | $N_I$ | 14797000 | [24] |
| | $ec$ | 9075189 | [25] |
| | $sp$ | 2904307 | [17] |
| | $ep$ | [0.062, 0.062, 0.126, 0.126, 0.126, 0.126, 0.126, 0.062, 0.062] | (A2),(A12) |

| | | | |
|---|---|---|---|
| **Brazil** | $P_i$ | [0.13679427,0.14659641,0.1562808667,0.1580315267,0.1344392667,0.11489118,0.07949242,0.04140749,0.01956642] | [4] |
| | $P_h$ | 0.01250014 | [13] |
| | $N$ | 213993441 | [26] |
| | $N_I$ | 6353000 | [27] |
| | $ec$ | 93951410 | [28] |
| | $sp$ | 21445651 | [19] |
| | $ep$ | [0.20，0.20，0.31，0.31，0.31，0.31，0.31，0.20，0.20] | (A2),(A12) |

[15]. U.S. Department of Homeland Security. Legal Immigration and Adjustment

of Status Report Fiscal Year 2021, Quarter 2. https://www.dhs.gov/immigration-statistics/special-reports/legal-immigration. Accessed 16 Oct 2021.

[25]. Our World in Data. South Africa: Coronavirus Pandemic Country Profile. https://ourworldindata.org/coronavirus/country/south-africa. Accessed 16 Oct

2021.

[26]. National Institute of Geography and Statistics of Brazil. Population. https://www.ibge.gov.br/en/home-eng.html. Accessed 16 Oct 2021.

[27]. Knoema. Brazil - International tourism, number of arrivals. https://knoema.com/atlas/Brazil/topics/Tourism/Key-Tourism-Indicators/Number-of-arrivals. Accessed 16 Oct 2021.

[28]. Our World in Data. Brazil: Coronavirus Pandemic Country Profile. https://ourworldindata.org/coronavirus/country/brazil. Accessed 16 Oct 2021.
